# Feasibility, acceptability, and sustainability of family-led postnatal care model: a multi-site mixed study in Ada’a District, Ethiopia

**DOI:** 10.1101/2025.03.11.25323725

**Authors:** Gadise Bekele Regassa, Konjit Wolde, Dedefo Teshite, Lello Amdissa, Walelegn Worku, Solomon Getachew Alem, Pooja Sripad, Anne Hyre, Stephanie Suhowatsky, Lisa Noguchi, Alemayehu Worku, Della Berhanu

## Abstract

**Objectives:** This study aims to assess the feasibility, acceptability, and sustainability of the family-led postnatal care model.

*Design:* The study applied a post-intervention mixed-method design drawing on cross-sectional survey, data review from registers and checklists, and in-depth interviews and key-informant interviews from February–April 2023.

*Setting:* The study was conducted in four health centers and catchment areas in Ada’a District, Oromia Region, Ethiopia.

*Participants:* The quantitative survey included 110 postnatal women. The qualitative components included in-depth interviews with postnatal women, husbands/partners, and family members and key-informant interviews with midwives/nurses, health extension workers, home care kit custodians, and health managers.

*Intervention:* Family-led postnatal care is a self-care innovation for postnatal women and newborns during their first week of life. In the model, a midwife or nurse invites family members to attend the discharge and assesses the mother and newborn using a pictorial checklist. The checklist is given to the families with guidance on retrieving a home care kit (containing blood pressure monitor, thermometer, and health-education booklet) from a volunteer community custodian. At home, families use the checklist and kit to assess the health of the postnatal mother and newborn for six days, returning the completed checklist and kit to the custodian afterwards.

*Main outcome measures:* The outcome measures are feasibility, acceptability, and sustainability of the family-led postnatal care model.

*Results:* Participants at facility and community levels felt that family-led postnatal care was feasible and acceptable due to the easy-to-use materials for varied literacy levels, its influence on spouses/partners and families to support mothers, and its empowerment of women to recognize signs that require care-seeking. All health centers continued the family-led postnatal care model, and most kits were functional six months after the end of project support.

*Conclusions:* Our results indicate that family-led postnatal care is a promising approach that can be tested and scaled in other settings.

*• Trial registration:* ClinicalTrials.gov (NCT05563974).

*Summary boxes:*

- **What is already known on this topic**

- In Ethiopia, 80% of mothers and newborns receive no postnatal care.
- Innovative models are needed to deliver postnatal care for mothers and newborns in Ethiopia.
- **What this study adds**

- This family-led postnatal care model’s culturally sensitive, low-literacy tools, and integration into the healthcare system have demonstrated significant feasibility, acceptability, and early signs of sustainability for a self-care approach to postnatal care in Ethiopia’s rural, resource-limited settings.
- **How this study might affect research, practice or policy**

- Family-led postnatal care, a self-care model for postnatal care, is a promising approach to test and scale in other settings.

## Background

In Ethiopia, maternal and newborn mortality are high at 412 deaths per 100,000 live births and 33 deaths per 1,000 live births, respectively.(1) These figures are significantly higher in rural areas where access to healthcare services is limited. The postnatal period, which encompasses the first six weeks after childbirth, is a critical time for both mother and newborn, accounting for up to 50% of maternal deaths and 40% of newborn deaths.(2) Despite this, only 17% of Ethiopian women receive postnatal care (PNC) within the recommended 48 hours after birth. This is largely due to cultural barriers that prevent women from leaving the house in the first weeks after delivery, lack of knowledge of the importance of PNC, perceived low quality of services, shortages in healthcare professionals, and inadequate infrastructure, especially in rural and resource-limited settings.(1,3–6) Consequently, innovative care models are urgently needed to improve PNC access and health outcomes.

The World Health Organization emphasizes the importance of locally tailored interventions, such as home-based postnatal visits, to monitor maternal and newborn health and enhance early detection of complications to initiate prompt seeking of care at the nearest health facility.(7) Different studies highlight that a combination of social support groups, community mobilization, and health worker training is crucial in overcoming cultural and systemic barriers to accessing PNC. These findings suggest that when community-driven innovations are integrated into PNC programs, maternal and newborn mortality can be reduced significantly in low-resource settings.(8–10)

Self-care practices have also emerged as a critical component in empowering mothers to manage their own health and that of their newborns during the postnatal period. The 2019 “WHO guideline on self-care interventions for health” outlines essential self-care practices such as exclusive breastfeeding, proper hygiene, nutritional management, and self-monitoring of postnatal symptoms.(11) In a study conducted in southwest Ethiopia, self-care practices were found to be inconsistent, with adherence rates as low as 30% for wound care and hygiene management due to socio-economic challenges, limited healthcare access, and cultural barriers.(12) Evidence suggests that family involvement in PNC can also reduce newborn mortality and increase maternal satisfaction with care.(13,14) These findings emphasize the need for more research to explore the role of self-care interventions in rural Ethiopian communities, particularly in overcoming the socio-economic and cultural barriers that limit access to PNC.

This study aimed to explore the feasibility, acceptability, effectiveness and sustainability of a family-led postnatal care (FPNC) model, an innovative model for PNC for mothers and newborns in Ethiopia. Effectiveness of the FPNC model in increasing PNC coverage has been reported separately (Regassa et al, 2024 under review). This paper focused on the feasibility and acceptability of the family-led model, which are critical in determining its sustainability and scalability. It emphasized the importance of evaluating the practicality of implementing such models within existing healthcare systems and community structures as essential.(15) Acceptability studies play a key role in ensuring that health interventions are culturally appropriate and well-received by the target population.(16) This study addressed these factors and aimed to provide insights that can inform the broader implementation of FPNC models in settings facing similar challenges.

## Methods

### Study design

The study applied a post-intervention mixed-method design drawing on a cross-sectional survey, data review from registers and checklists, and in-depth interviews and key-informant interviews from February to April 2023.

### Study setting

This study took place in four health centers and their catchment areas in Ada’a District, Oromia Region, Ethiopia. Ada’a is one of the districts in East Showa zone, where most of the population resides in rural areas. In Ethiopia, districts’ primary health care system consists of a primary hospital, five health centers, and five health posts under each health center. Health extension workers (HEWs) provide preventive and curative services, including maternal, newborn, and child health services, at health posts and through home visits. Deliveries are expected to take place in health centers and hospitals.

Four health centers in Ada’a District were purposively selected to participate, and all surrounding communities were eligible to participate. One health center was involved in the conceptualization, design, and beta testing of the FPNC model components and was excluded from the study.

### Description of the FPNC intervention

Human-centered design activities revealed key barriers to timely PNC use in the local context that informed intervention design. Mothers and newborns were expected to stay home during the first 42 days, particularly during the first 10 days after birth. Mothers relied mainly on their husbands/partners, immediate family, religious leaders, and neighbors for advice and decision-making. They often felt pressured to “prove” themselves to be capable mothers and feared being inadequate at caring for their newborn or recognizing problems. Health facilities were typically seen as a last resort, sought only in critical situations.(17)

These insights informed the FPNC intervention, a novel model for reaching postnatal women and newborns during the first week after birth with key PNC services, founded on self-care principles. FPNC consists of the components described in Figure 1.

**Figure 1.**
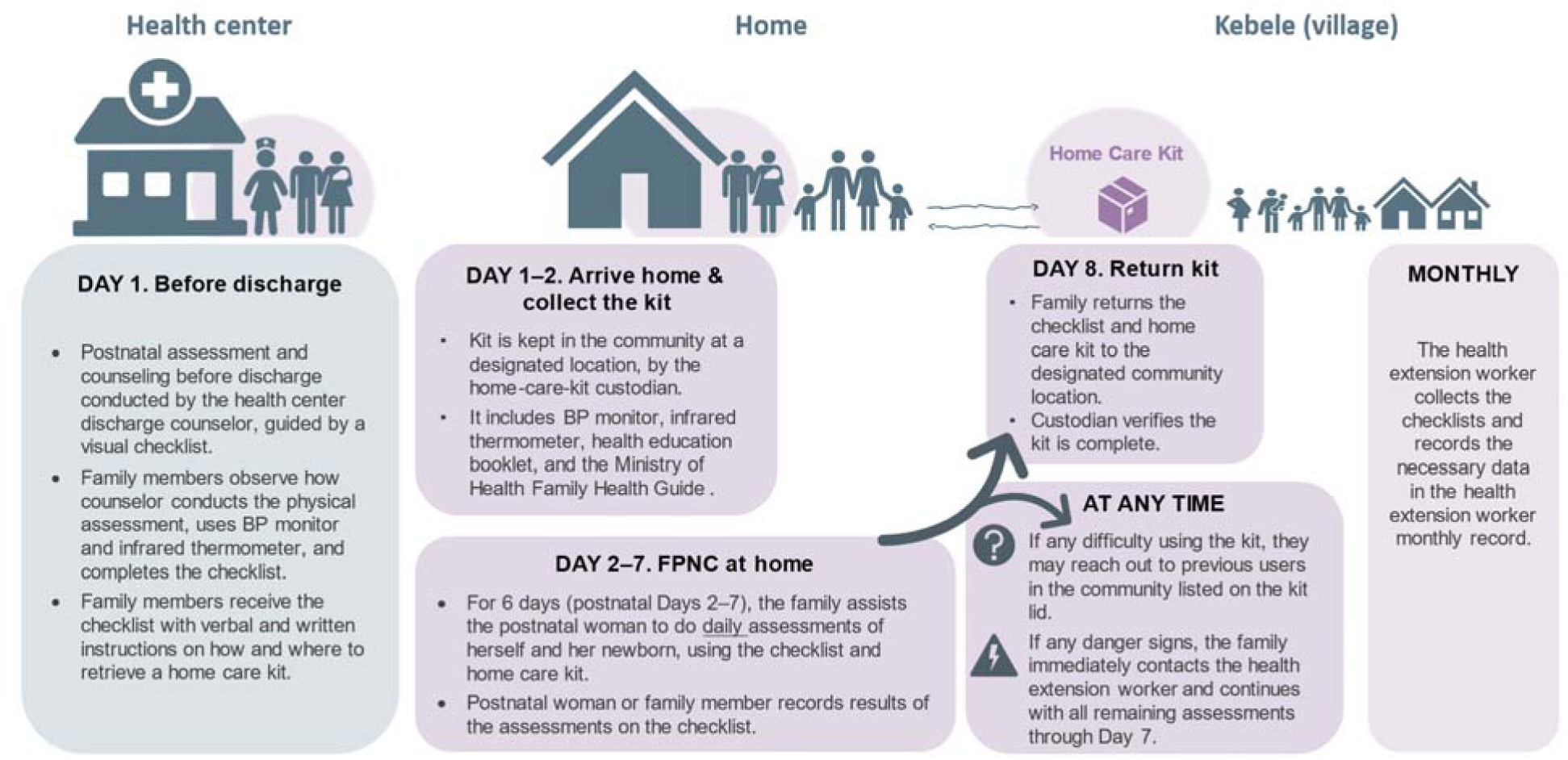
Implementation of the FPNC intervention.

The intervention included training midwives, nurses, and health officers to equip them with skills to provide pre-discharge care effectively as part of FPNC. The study team developed a script for the healthcare providers to use alongside a checklist during discharge, ensuring consistent communication and assessments. The checklist included manual assessments of mothers and newborns and assessments using devices such as a thermometer and blood pressure (BP) monitor, which were made available to families in the community as a home-care kit (HCK).

The study team also developed a photo booklet, which was included in the HCK, that featured images of normal and abnormal signs in mothers and newborns to aid caregivers in identifying potential issues. The HCK itself was a portable case, labeled with a sticker containing contact information for previous users, allowing current users to seek guidance if needed.

The study team along with the HEW and community leaders designated custodians to manage the kits, based on specific criteria. The custodians maintained a register to record who borrowed the HCK and returned it with the checklist. They also inspected the kits upon return, recharging any components as necessary.

### Patient and public involvement

We collaborated with a midwife at one health center to develop the checklist and identify the necessary equipment for the homecare kit. In partnership with health extension workers and community leaders, we identified community custodians. Additionally, we worked closely with these custodians to establish a mechanism for tracking and managing the homecare kits.

We engaged the district office to introduce the intervention, health centers to oversee the distribution of homecare kits, and health extension workers to manage data collection using the checklist. Finally, we invited participants and community members to a results dissemination meeting.

### Sample size

#### Quantitative sample

The sample size was calculated using the 2016 Ethiopian Demographic and Health Survey coverage of PNC within 24 hours of 17%.(1) With a desired increase to 45% due to the intervention, a 5% level of significance, 80% power, a design effect of 2.0, and a non-response rate of 10%, the minimum sample size required to measure our primary outcome was 109 postnatal women at pre- and post-intervention.(1) This paper focuses on the post-intervention survey only. The quantitative sample included postnatal women who gave birth at the four study health centers; women were included sequentially until the sample size was met.

#### Qualitative sample

Forty key-informant interviews and 48 in-depth interviews were conducted. The qualitative sample included: 1) eligible women who gave birth at the four study health centers; 2) eligible partners; 3) eligible women’s family members; 4) discharge counselors at the four health centers; 5) HEWs in the catchment area of the four health centers; 6) HCK custodians in the community; and 7) district health managers and health center managers. All participants were a minimum of 15 years old. Purposive sampling was employed, based on the completed postnatal checklists and consultation with HEWs and the HCK custodians (Table 1).

**Table 1.** Inclusion criteria of the study participants for qualitative data collection.

| Study population | Description |
| --- | --- |
| Health center discharge counselors | All eligible discharge counselors on staff of intervention health center who were midwives, nurses, and/or health officers |
| Postnatal women | Selected eligible women who gave birth at intervention health centers (women who had a postnatal check on Day 6 after delivery, women who missed a postnatal check, women who identified postnatal danger signs and sought care from a health provider, women who identified postnatal danger signs but who did not seek care) |
| Family members | Family members of eligible women present at discharge |
| Husbands/partners | Husbands/partners of eligible women |
| HEWs | All HEWs working in catchment area of intervention health centers |
| HCK custodians | Community leaders serving as HCK custodians in catchment area of intervention health centers |
| Health managers | Health managers overseeing maternal/newborn health activities in Ada'a District; heads of intervention health centers |

### Data collection procedure

#### Quantitative

Training sessions were conducted for field data collectors and supervisors, focusing on study objectives, procedures, and ethical considerations. A comprehensive survey manual of standard operating procedures was developed and used during the training, pilot phase, and survey execution. The training also covered the appropriate use of tablets for data entry during data collection.

Health center staff responsible for postnatal discharge at the four selected centers were tasked with screening mothers for study eligibility. Before discharge, eligible mothers were asked if they were interested in learning more about the study. If they expressed interest, they were referred to FPNC study staff. Study personnel on-site approached the mothers and their family members, provided further details about the study, addressed any questions, and obtained oral informed consent. For those who consented, contact information was collected and passed on to the local study enumerators. Designated data collectors visited the mothers’ homes on postnatal Day 8 to conduct surveys.

#### Qualitative

HEWs and HCK custodians were invited to participate in community orientations. After the post-intervention quantitative survey, research assistants approached health center discharge counselors; postnatal women and family members, husbands/partners; HEWs; and HCK custodians at a convenient place, explained the study, and obtained oral informed consent before they participated in the qualitative data collection.

In-depth interviews were conducted with postnatal women at their homes who met the description outlined in Table 1. Key-informant interviews were conducted with the discharge counselors, HEWs, health managers, and HCK custodians who were involved in FPNC intervention implementation. The interviews were conducted at their place of work. Each interview was audio recorded with the permission of the participants to ensure accurate data capture.

### Outcome measures

#### Feasibility

- Feasibility of FPNC at health center, community and at household level
- Proportion of women discharged from the facility whose family member retrieved the HCK, proportion of women who report successful use of BP cuff, proportion of women who report successful use of thermometer, proportion of HCKs returned by Day 8, proportion of checklists returned with the HCK, proportion of checklists collected by HEW

#### Acceptability

- Proportion of women who give birth at health facility and accept a checklist from the discharge counselor, proportion of women who report they prefer this PNC approach, proportion of women who report confidence in this PNC approach
- Perception of HCK custodians, health managers, midwives/nurses, and HEWs regarding the FPNC approach
- Perception of woman’s partner and family regarding the FPNC approach

#### Sustainability

- Continued implementation of the FPNC model at the health centers and communities six months after the post intervention survey was completed.

### Data management and analysis

Quantitative data were collected electronically using SurveyCTO application on tablets. Data from the field were synchronized daily to the central cloud server. Study team leads conducted daily checks on completeness and consistency of the collected data. The electronic data capturing system had built-in logic, range, and skip patterns to limit data inaccuracies. Data were cleaned before the main analysis. Descriptive analysis using frequency and proportion was computed. STATA version 15 software was used to analyze the data.

Qualitative data were interpreted using thematic analysis to understand participants’ experiences with FPNC and thoughts on its feasibility and sustainability. The study team actively constructed themes and derived and patterns (or meanings) collectively. Themes were generated by the study team familiarizing themselves with the data, generating initial codes, searching for themes, reviewing themes, and defining and naming themes through a deliberative process. Data were transcribed and translated to English. Open code software was used for analyzing the qualitative data.(18)

### Ethical approval

The study obtained ethical approval from the Johns Hopkins Bloomberg School of Public Health Institutional Review Board (IRB No. 21096) and the Addis Continental Institute of Public Health Institutional Ethical Review Board (IRB No. 0029). All participants gave informed consent before taking part in the study.

## Results

### Characteristics of postnatal women

More than 90% of the postnatal women in the post-intervention survey were 18 to 35 years old. Three-quarters had no education or a primary education, and 24% had secondary or above. Orthodox religion was the predominant religion (87%). More than 90% of interviewed women were either currently married or cohabiting (Table 2).

**Table 2.** Background characteristics of women included in post-intervention surveys (February–April 2023), Ada’a District, Ethiopia.

| Variable | N = 110 |
| --- | --- |
|  | n (%) |
| **Age in years** |  |
| 15–24 | 45 (41) |
| 25–34 | 49 (45) |
| ≥ 35 | 16 (14) |
| **Education level** |  |
| None | 26 (24) |
| Primary | 58 (52) |
| Secondary and above | 26 (24) |
| **Religion** |  |
| Orthodox | 96 (87) |
| Other | 14 (13) |
| **Marital status** |  |
| Currently married/cohabiting | 103 (94) |
| Other | 7 (6) |

### Feasibility: At health center

The major themes that emerged from analysis of the key-informant and in-depth interviews under feasibility included the feasibility of implementing the FPNC model at the health center, community, and household levels.

Most discharge counselors managed to follow the scripts during discharge counseling, although they stated that the counseling focused more on how to use the BP and thermometer devices than on other danger signs. Regarding adequacy of the information and checks for detecting danger signs in the mother and newborn, they all agreed that the materials were enough and were also feasible for individuals with minimal or no education. However, some raised the concern that there was only one midwife covering antenatal care, delivery, and PNC, which resulted in increased workloads when doing the discharge counseling. They also stated that it took providers more time to counsel families with minimal or no education.

Health managers agreed that the FPNC approach had made the postnatal discharge systematic and effective because of the script and checklist provided. A health manager at a woreda stated:

> *“In the past, the healthcare provider might have missed important details while providing the service because they lacked an organized checklist form, but now that FPNC has provided the checklist, we follow it, which helps the healthcare provider to provide discharge-counseling services at the required level.”*

The discharge counseling also included family members. As such, postnatal women and family members were happy to have family members present during counseling. A mother explains:

> *“Very good, being with the family first makes my family understand it too, because that advice is very important to me and the baby, and my family can do the same help for me and the baby that the health worker did for us. It’s very good for me.”*

A husband commented on the discharge counseling:

> *“It’s good because knowing what’s going to happen beforehand means there’s nothing to panic about. Because if something happens, they said come to us [the health center].”*

### Feasibility: At community or community

Ninety-three percent of the women reported no difficulties in receiving and returning the HCK to the custodian. Among those who reported a challenge, the most common challenges were that the HCK custodian was not available, there were no HCKs available, and the postnatal woman did not have a family member available to retrieve the HCK. Data abstracted from the HCK registers shows that 100% of the HCK and 98% of the checklist were returned with the HCKs (Table 3).

**Table 3.** Feasibility of retrieving and returning the HCKs through the FPNC approach, reported in post-intervention surveys (February–April 2023), Ada’a District, Ethiopia.

| Variable | N = 110 |
| --- | --- |
|  | n (%) |
| **Challenges with obtaining the HCK** |  |
| Had no challenges in receiving the HCK from the custodian | 99 (90) |
| The HCK custodians were not available | 4 (4) |
| There were no HCKs available | 3 (3) |
| I didn’t have a family member available to retrieve the HCK | 3 (3) |
| No challenges in returning the HCK to the custodian | 102 (93) |
| **Data extracted from HCK register** | **N = 99\*** |
| HCKs returned | 99 (100) |
| Checklists returned with the HCKs | 97 (98) |
| Checklists collected by HEWs | 73 (74) |

\* Data is for 99 women registered on the HCK register.

In qualitative interviews, the HCK custodians reported being comfortable in filling the register. Only one mentioned having encountered issues in filling the form due to difficulty in understanding the language in the form. Overall, most custodians were happy with their new role. One HCK custodian stated:

> *“I am happy in my work because I want mothers to not face any problems, and I am also a mother, so I am happy to be chosen for this service. If there is anything else, I will do it, and it is going well. I haven’t had any problems yet.”*

Some stated that their role as custodian interfered with their daily activity. One custodian raised the challenge she had encountered:

> *“When I am working on something, sometimes it interrupts me and takes time when checking and giving the materials.”*

To facilitate availability of the HCKs in their absence, some custodians delegated other family members to carry out their role thereby reducing the chance of a family not getting the kit. HEWs appreciated the role of HCK custodians as they helped keep the kits safe.

About 73% of the returned checklists were collected by the HEW (Table 3). Some of the HEWs stated that collecting the checklist from the custodians was a challenge. One HEW stated:

> *“Whenever we went, we checked all their checklists, materials, what is available or not, who took if their signatures were there, saw how many checklists and for whom they were given, collected, and brought that here.”*

### Feasibility: At home

Most families did not face challenges while using the HCK. When there was a problem, non-functioning BP monitors and thermometers were the most common challenges encountered. In contrast, only a few women (38.2%) used the photo health-education booklet. Among those who used it, nearly all reported not encountering any challenge in its use (Table 4).

**Table 4.**
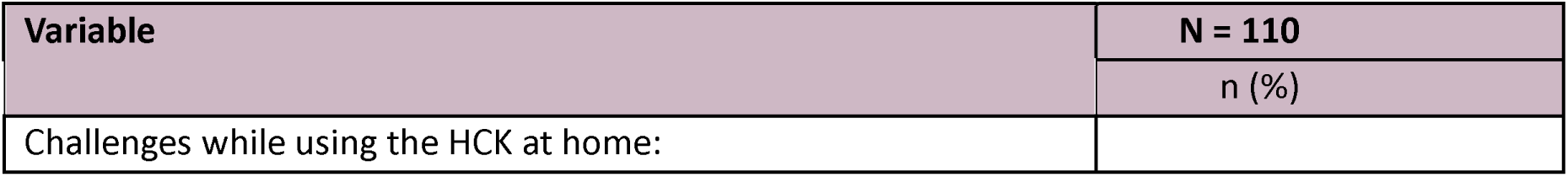
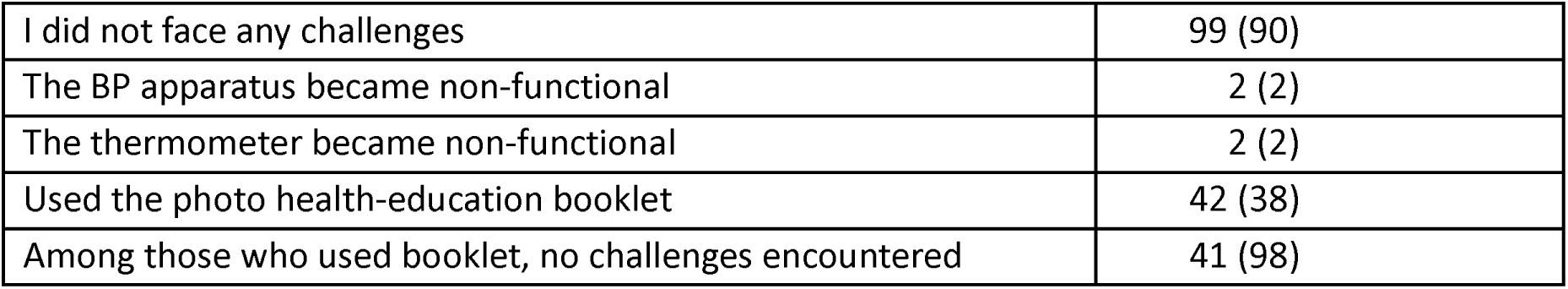
Feasibility of using the HCK, reported in post-intervention surveys (February–April 2023), Ada’a District, Ethiopia.

More than 80% of the women used the BP device, thermometer, and checklist without difficulty and only less than 14% of women had difficulty (Figure 2). In qualitative interviews, women were able to use the checklist and devices and even most of the least educated women reported using the contents of the HCK. A postnatal woman explains how she used the kit:

> *“I am illiterate myself and I am confident I can use the checklist and devices for myself and even for others too because of the advice given at the health center and I was also able to understand the pictorial display.”*

**Figure 2.**
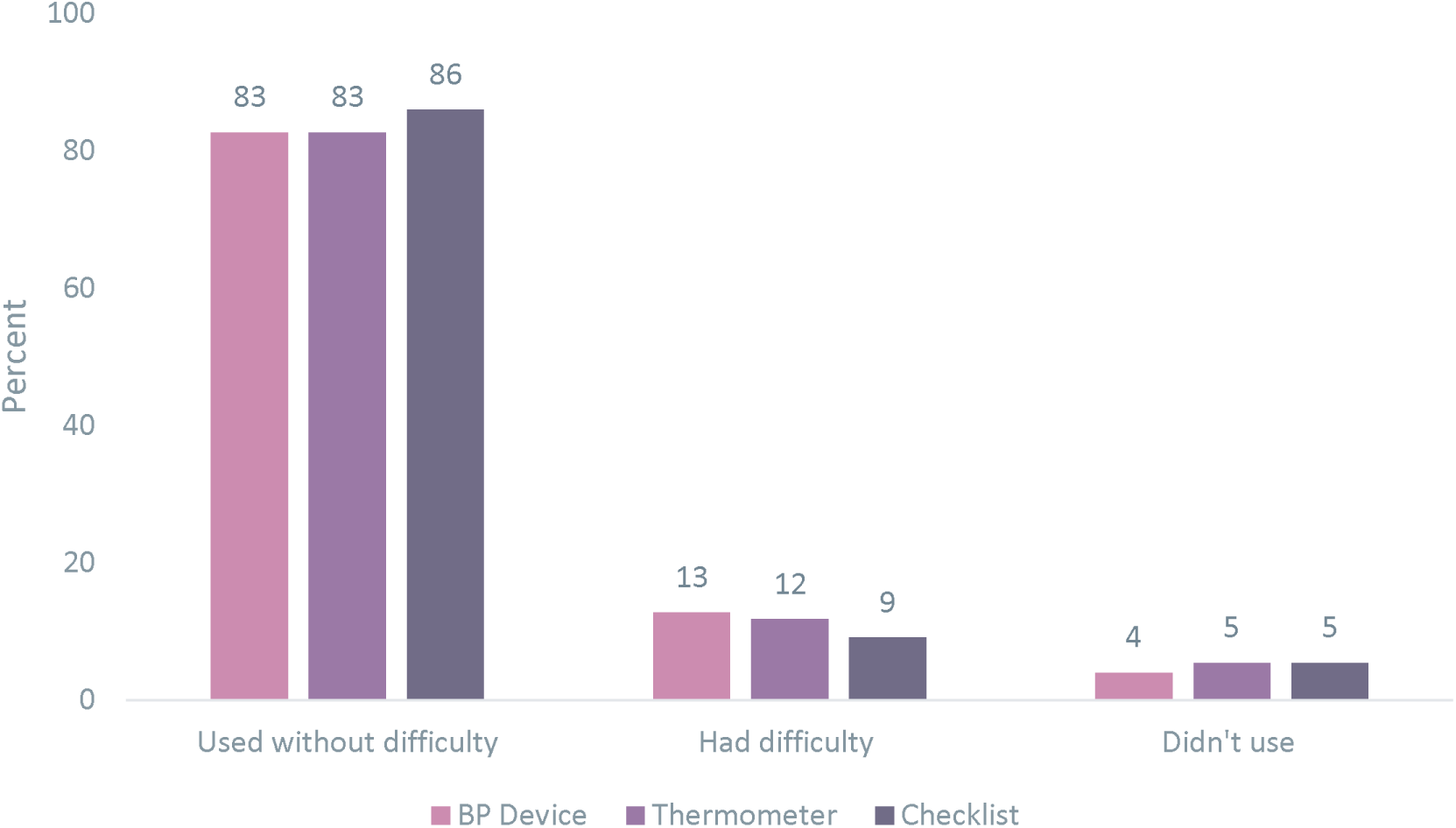
Feasibility of postnatal women and their families using the BP monitor, thermometer, and checklist during the post-intervention period (February–April 2023) N=110, Ada’a District, Ethiopia.

However, the checks were more focused on the devices while other danger signs without the devices were given less attention. Women and family members who had not done the recommended assessments thought that the devices could assess the remaining checklist items. A mother stated the following:

> *“I was sure, because they explained to us that if there is a problem, if there is pain, it is the device that tells you without speaking, if it lights up red, I will say there is a problem.”*

Per Ethiopian national guidelines, HEWs are expected to provide home visits on Day 1, 3, and 7 post-delivery. For the HEWs who did their home visits, they reported that most of the FPNC families managed to use the kit and checklist properly. HEWs stated that although some families contacted them to solve challenges encountered on how to use the checklist and kit (BP cuff and thermometer), they highlighted that the custodians were normally the first ones to be contacted when families encountered a challenge. HEWs recommended giving more training to empower the HCK custodians as they are closer to the community.

### Acceptability

Acceptability themes included postnatal women’s and their families’ self-care experience with the FPNC approach and the acceptability of the FPNC model by healthcare providers and custodians.

Out of 110 women who were discharged with a checklist, 105 (95.5%) retrieved the HCK from the custodians in their community. Most of the women (94.5%) preferred the FPNC approach compared with the traditional PNC by a healthcare provider during a previous childbirth experience. Ninety-five percent of the women felt confident that they, along with their babies, had received good quality PNC when using the HCK at home. Almost all women wanted to use the FPNC approach for future births. Women reported that husbands/partners were “very involved” in the FPNC model (Table 5).

**Table 5.** Acceptability of the FPNC approach in the post-intervention period (February–April 2023), Ada’a District, Ethiopia.

| Variable | N = 110 |
| --- | --- |
|  | n (%) |
| Obtained the HCK after discharge | 105 (96) |
| **PNC approach preference compared to previous birth** |  |
| Prefer traditional PNC by a healthcare provider | 2 (2) |
| Prefer FPNC | 104 (95) |
| No previous PNC for comparison | 4 (3) |
| Confident in the quality of PNC when using HCK | 104 (95) |
| **Level of confidence in the quality of assessments using the HCK** |  |
| Highly confident | 98 (89) |
| Moderately confident | 11 (10) |
| Not sure or do not know | 1 (1) |
| **PNC preference for future birth** |  |
| FPNC home-based approach | 107 (97) |
| PNC by a HEW or at health center | 3 (3) |
| **Husband or partner involvement in the FPNC care** |  |
| Very involved | 96 (87) |
| Somewhat involved | 8 (7) |
| Not involved | 6 (6) |

### Self-care experience of postnatal women and their families

Women’s and families’ acceptance of FPNC was further described through the agency they felt using the self-care approach. The checks for the postnatal women were done by family members, and the newborn checks were mainly done by their mothers, the postnatal women. Postnatal women reported being pleased with the help they received from their husbands/partners and family members to conduct the health checks at home. Postnatal women described what they liked:

> *“When they [family members] started taking my temperature and blood pressure regularly at home, I was especially happy. I appreciate the information that was given to us.”*
>
> *“I am very happy with my health because I will not make my families worried about my health and I myself will not be worried as well. Because this one [FPNC] is here to help me, this item is not for someone else. I am the one who explains to my husband when it [baby] has a problem or if I am sick. He will help me by bringing and checking using the item and then he will understand whether it is certain or not. He has faith in it, though nothing has happened to me, I am happy with it.”*
>
> *“When people offer their help and support, it is a demonstration of their care and love for you. It’s wonderful to have loved ones who are there for you in times of need.”*

Husbands also were happy and appreciated that they were trusted to follow and actively support the health of their wife/partner and newborn. They reported that with FPNC their engagement was different from previous births when they were not as actively involved, in part due to sociocultural norms. A husband describes how he was more engaged in the postnatal period:

> *“I will take care of my baby before anyone else. As per tradition, males are not allowed to go inside where the mother who delivered is resting, and they never eat what is prepared for the mother. But now, I take care of both the mother and the baby.”*

Another husband explained:

> *“Previously, with our tradition … there wasn’t much we understood until they got very sick. But now we have understood a little because of this. We have understood that it is very useful to have health follow-up even when the kit is not brought. There are those who never even go, delivering at home. But for us now … the other children were born at home; now I have the awareness that there is follow-up after delivery, before delivery. The awareness we found after delivery gave us freedom, that’s it.”*

Husbands/partners were the most involved family members in the FPNC approach and their preferred roles in FPNC were most corroborated by women. Husbands were the most preferred, was followed by the postnatal women’s mothers, to use the devices to perform health checks (Figure 3). In addition, older children, mothers-in-laws, and sisters provided checks to mothers and newborns. An eldest son describes what his role was in the FPNC:

> *“We assisted her in maintaining her hygiene, and I have been taking the newborn’s and mother’s blood pressure and temperature. I’ve been filling the checklist.”*

**Figure 3.**
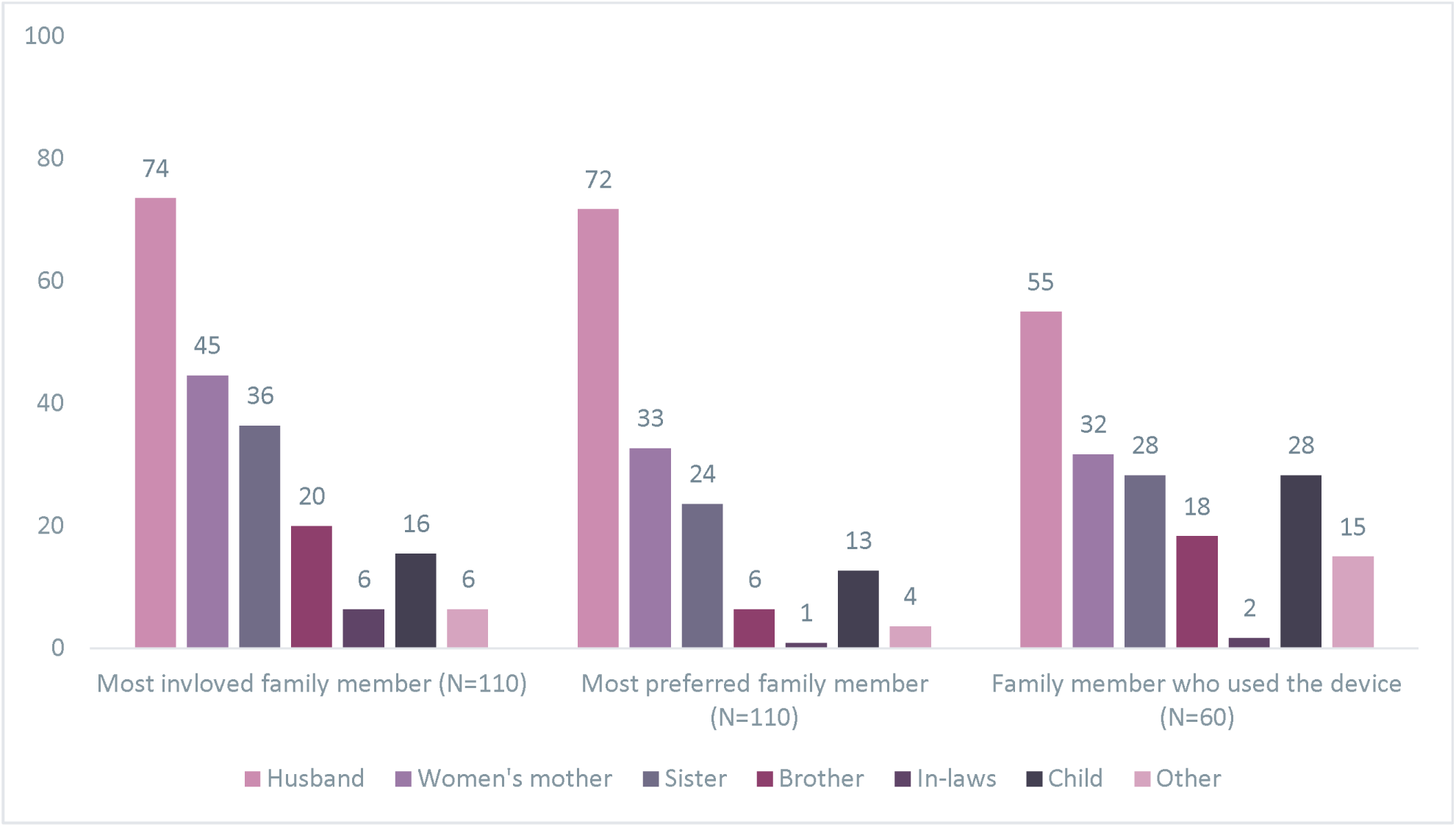
Family involvement and use of devices in the post-intervention period (February–April 2023), Ada’a District, Ethiopia.

### Healthcare providers’ and managers’ attitudes towards the FPNC approach

Discharge counselors had a positive attitude towards the FPNC approach stating it benefited the community, helped women take care of their health, and contributed to maternal health-seeking behavior. An HCK custodian explained:

> *“I’m happy that mothers are getting help even at home. Mothers used to deliver at home and bleed, but now they deliver at health facilities and get the follow-up at home too. When they see red, they go to the health facility.”*

Most HEWs stated that FPNC has made things easier for them because the women learned to care for themselves with their families’ help. HEWs reported that women, families, and communities have an increased awareness to do more health checks, especially BP measurements. Family members and women had contacted them upon identifying danger signs. The HEWs stated that when families expressed concerns around identified danger signs, they were appropriate and HEWs managed them accordingly.

Most managers agreed that the FPNC approach has made it possible for many women to assess their health in the postnatal period at home. They also implied that early recognition of postnatal problems and follow-up by HEWs has become easier since the family and postnatal women were doing the checks on a daily basis. A health manager explained why he supported the approach:

> *“Yes, I support [FPNC], because people might get hurt when they hesitate to come to health center thinking that they might get better after a day or after. Before the FPNC, only the mother used to be counseled but now the whole family gets the counseling, so the attention increased and also the equipment [to identify a danger sign] alarms them.”*

### Sustainability of FPNC approach

All four health centers were still providing FPNC services six months after the post intervention survey was completed, as reported in October 2023. Among 64 HCKs provided to custodians, 62 (96.7%) were functional.

Sustainability themes considered the health managers’ perspectives, included their commitment to and challenges around the HCK. All health managers confirmed their commitment to sustaining FPNC as they saw the benefits to the women and their communities. They recommended integrating the FPNC approach into monthly and quarterly reports to increase accountability and responsibility for monitoring the approach’s progress. They also mentioned that issues related to the resource supply and kit maintenance and replacement might pose a challenge to FPNC’s sustainability. A health manager described the sustainability of FPNC this way:

> *“It needs attention from higher levels of government, woreda, and community. This needs supplies so it needs higher level involvement. There are procedures for ANC [antenatal care], PNC, and others that are set by the government, so it would be good if this has such procedures from the government. If training is given and the supplies are provided, I believe this can work.”*

## Discussion

This study used a mixed-methods approach to comprehensively evaluate the acceptability, feasibility, and sustainability of the FPNC model, drawing on the perspectives of diverse stakeholders to capture a holistic understanding. The FPNC model was implemented across four health centers in the Ada’a District, Oromia Region, Ethiopia. The self-care model demonstrated high feasibility and acceptability, particularly due to the simplicity and accessibility of the materials provided, which were designed for varied literacy levels. At both the facility and community levels, participants reported that the intervention positively influenced families—particularly husbands/partners—by engaging them to support mothers during the postnatal period. Many women reported increased confidence in recognizing danger signs that required urgent medical attention. This aligns with a broader finding that culturally appropriate interventions tend to achieve higher rates of success via self-care mechanisms.(12) After six months, all health centers continued to use the FPNC model, and almost all of the HCKs remained functional and in use.

The FPNC model aimed to address the extremely low coverage of PNC in Ethiopia.(1, 19) Regassa GB et al. (unpublished observations) reported that the FPNC intervention was effectiveness at significantly increasing PNC coverage. Before the intervention, fewer than 11% of mothers and newborns received a postnatal check on Day 3 and Day 7 after birth. However, post-intervention, this figure rose to over 95%, indicating notable progress in having timely PNC. Both this paper and the one evaluating FPNC’s effectiveness highlighted that very few women received PNC from an HEW, either at home or in a health facility.

The acceptability and feasibility of the FPNC model largely stem from its alignment with cultural norms. In Ethiopia and sub-Saharan Africa, where postnatal women traditionally remain at home for recovery, the model’s cultural fit increased acceptance.(17,20–22) In southwestern Ethiopia, 75% of women adhered better to PNC when cultural expectations were respected.(12) Similar interventions in Uganda and Zambia boosted healthcare engagement by up to 20%.(23,24) The involvement of respected community figures improved health outcomes and raised postnatal care coverage by 25%–30%.(8,25) In our study, though custodians expressed concerns about workload, they maintained their roles six months post-intervention.

Our findings indicated that husbands/partners and family members were involved in the postnatal discharge at health centers and checks done at home. Studies show that inviting and involving husbands/partners and family members in maternal and newborn clinics has resulted in higher engagement in the health of the women and newborns.(13,14,26–28) Involving husbands/partners and families in discharge counseling might have resulted in higher engagement following the women and newborn health after birth. Future research on self-care models, such as FPNC, may consider exploring possible long-term effects of involving husbands/partners and families on equitable gender and power dynamics in household decision-making around health.

Despite 28% of the women in the study having no formal education and living in rural areas with typically low literacy levels, the women’s capability to engage in self-care using HCK proved highly feasible. This is consistent with other findings that have reported that visual aids and simplified instructions can bridge literacy gaps in low-resource settings.(29) Some women and families did misunderstand the requirements for completing all necessary checks, assuming that the devices covered everything. These findings underscore the need to ensure that the information provided to families at discharge is clear and that it emphasizes the importance of performing all recommended checks appropriately.

Healthcare providers involved in FPNC embraced the self-care-prompting model. By incorporating structured scripts and visual aids to guide postnatal counseling, the model ensured that health professionals could offer standardized, comprehensive advice without overburdening their workloads.(29,30) A few did mention that counseling with the level of detail that the script and checklist required took more time.

Six months after the intervention, 100% of health centers involved in the study continued to implement the FPNC model, and nearly all HCKs were still in use. This reflects a strong level of sustainability, which has been linked to community ownership and system integration. A systematic review found that maternal health programs in sub-Saharan Africa were more likely to be sustainable when communities feel a sense of ownership and when interventions are integrated into existing healthcare systems.(31) In this case, the FPNC model’s integration into the local healthcare system and its adaptability to community needs facilitated its continued use, suggesting the potential for broader scalability. However, the short follow-up period of six months limits the understanding of long-term sustainability.

This was a small study so the findings might not be generalizable to other settings. Because all the women in this study delivered in a facility, we did not assess the acceptability or feasibility of the model for home deliveries, which remain common in Ethiopia, with rates as high as 49%.(32) No simultaneous control, potential for confounding by time, and self-reported measures can be subject to bias. Future research should evaluate FPNC in a rigorous randomized controlled trial in Ethiopia and other settings, as well as feasibility among families with home births.

## Conclusion

The FPNC model’s culturally sensitive, low-literacy tools, and integration into the healthcare system have demonstrated significant feasibility, acceptability, and early signs of sustainability for a self-care approach to PNC in Ethiopia’s rural, resource-limited settings. These findings align with broader global health findings, showing that tailored, community-centered self-care interventions can significantly enhance PNC delivery, even in the most challenging environments.

## Data Availability

The de-identified data that support the findings of this study are available on figshare using this DOI 10.6084/m9.figshare.28572035.

https://figshare.com/s/bd3cf186b83e74a0c59b

## Transparency

The lead author (GBR) affirm that the manuscript is an honest, accurate, and transparent account of the study being reported; that no important aspects of the study have been omitted; and that any discrepancies from the study as planned (and, if relevant, registered) have been explained.

## Authors’ contributions

GBR, KW, DT, LA, WW, AH, SS, LN, AW, and DB contributed to the conceptualization of the study. GBR and AW analyzed and interpreted the data. GBR drafted the manuscript. All authors contributed and guided the analysis and writing of the manuscript. All authors have read and approved the final manuscript.

The corresponding author attests that all listed authors meet authorship criteria and that no others meeting the criteria have been omitted.

